# Mucosal-Associated Invariant T (MAIT) Cells are Highly Activated in Duodenal Tissue of Humans with *Vibrio cholerae* O1 Infection

**DOI:** 10.1101/2021.06.17.21258781

**Authors:** Taufiqur R. Bhuiyan, M. Arifur Rahman, Shubhanshi Trivedi, Taliman Afroz, Hasan Al Banna, Mohammad Rubel Hoq, Ioana Pop, Owen Jensen, Rasheduzzaman Rashu, Muhammad Ikhtear Uddin, Motaher Hossain, Ashraful I. Khan, Fahima Chowdhury, Jason B. Harris, Stephen B. Calderwood, Edward T. Ryan, Firdausi Qadri, Daniel T. Leung

## Abstract

Mucosal-associated invariant T (MAIT) cells are unconventional T lymphocytes with a semi-conserved TCRα, activated by the presentation of vitamin B metabolites by the MHC-I related protein, MR1, and with diverse innate and adaptive effector functions. The role of MAIT cells in acute intestinal infections, especially at the mucosal level, is not well known. Here, we analyzed the presence and phenotype of MAIT cells in duodenal biopsies and paired peripheral blood samples, in patients during and after culture-confirmed *Vibrio cholerae* O1 infection. Immunohistochemical staining of duodenal biopsies from cholera patients identified MAIT cells in the lamina propria, but not in the lining of villous and crypt epithelia. We observed significantly higher frequencies of duodenal MAIT cells at the acute stage (day 2) of *V. cholerae* infection as compared to late convalescence (day 30, *p* = 0.0049). By flow cytometry, we showed that duodenal MAIT cells are more activated than peripheral MAIT cells (p < 0.01 across time points), although there were no significant differences between duodenal MAITs at day 2 and day 30. We found fecal markers of intestinal permeability and inflammation to be correlated with the loss of duodenal (but not peripheral) MAIT cells, and single-cell sequencing revealed differing T cell receptor usage between the duodenal and peripheral blood MAIT cells. In summary, we show that MAIT cells are present and highly activated in the lamina propria of the duodenum during *V. cholerae* infection. Future work into the trafficking and tissue-resident function of MAIT cells is warranted.

## Introduction

Mucosal-associated invariant T (MAIT) cells are a recently identified non-conventional T cell subset. They express an invariant T cell receptor (TCR) Vα chain (Vα7.2 or TRAV1-2 in humans) and a variable but restricted number of TCRβ chains. MAIT cells are found in mucosal tissues and associated organs, including as the liver, lung, mesenteric lymph nodes, and intestinal epithelium [1]. In human peripheral blood, MAIT cells constitute approximately 1-10% of total T lymphocytes [2], and they account for up to 40% of T cells in the liver [3]. In the human intestine, they are located in both the lamina propria and as part of the intraepithelial lymphocyte compartment [4]. The ligand for MAIT cells has been identified as belonging to a class of transitory intermediates of the riboflavin synthesis pathway [5], which are produced by many, but not all, bacteria and yeast. These vitamin B metabolites are presented on the surface of MR1 [6], the non-polymorphic MHC class I related protein. MAIT cells are capable of releasing IFN-γ, TNF-α, and IL-17 in response to stimulation, and they also possess cytotoxic activity [7, 8], killing infected cells via granzyme B and perforin.

Cholera is a life-threatening diarrheal disease caused primarily by *Vibrio cholerae* O1, responsible for close to 3 million cases and 100,000 deaths annually in endemic countries alone [9]. The mechanisms of protection against cholera are not well understood, although we have previously shown that in patients hospitalized with severe cholera, both adaptive and innate immune responses are induced. We have demonstrated increases in circulating *V. cholerae* antigen-specific antibodies, as well as antigen-specific memory B and memory T responses in both children and adults [10, 11]. We have also shown, through endoscopically-obtained duodenal biopsies, that there is an increase in cells of the innate immune system and their mediators during acute cholera [12, 13].

We have previously reported that circulating MAIT cells are activated during *V. cholera* O1 infection, and that in children, but not adults, their circulating numbers are significantly decreased by day 7 after infection and onward [14]. We also demonstrated an association between circulating MAIT cells and class-switched antibody responses against LPS. Despite their abundance in the intestinal mucosa, little is known regarding the activity of MAIT cells in mucosal tissue during acute enteric infection. Thus, our objective was to characterize MAIT cells in the intestinal mucosa during *V. cholera* infection.

## Methods

### Study population and sample collection

We enrolled patients admitted to the Dhaka Hospital of the International Centre for Diarrhoeal Disease Research, Bangladesh (icddr,b) who had acute watery diarrhea and positive stool cultures for *V. cholerae* O1. We recruited patients who had no underlying medical conditions and had an otherwise normal physical examination and baseline laboratory parameters. All patients were rehydrated and provided antibiotics per hospital protocol prior to enrollment and were hemodynamically stable at the time of the procedure. After informed consent, we performed esophagogastroduodenoscopy (EGD) on day 2 (acute illness phase) and day 30 (convalescent phase) following admission. Using standard forceps, we obtained approximately six duodenal pinch biopsies of approximately 1 mm^3^ in diameter from each patient at each time point. We also obtained a stool sample from each patient at day 2 and venous blood samples at days 2 and 30.

### Phenotyping of MAIT cells by flow cytometry

From venous blood samples, we isolated peripheral blood mononuclear cells (PBMCs) by differential centrifugation on Ficoll-Isopaque (Pharmacia, Piscataway, NJ). We stored plasma at −80°C for use for immunological assays as detailed below. From duodenal biopsy samples, we isolated lamina propria lymphocytes (LPLs) by incubation in 1 mM EDTA and 1 mM dithiothreitol (DTT) followed by filtering through a nylon cell strainer and treatment with collagenase and DNase, as we have previously described [15]. We washed and stained the freshly isolated PBMCs and LPLs with an established fluorochrome-conjugated antibody panel designed for MAIT cell isolation. Antibodies were purchased from BioLegend (San Diego, CA), BD Biosciences (San Jose, CA), or Life Technologies (Carlsbad, CA): Vα7.2-PE, CD3-PE-Texas Red, CD4-Amcyan, CD8-FITC, CD161-APC, CD38-PE-Cy7, CD69-PerCP-Cy5.5 and DAPI. After 45 minutes incubation at 4°C, we analyzed at least 10^5^ lymphocytes on a FACSAria III flow cytometer (BD Biosciences, San Jose, CA) and analyzed data using FlowJo 10 software (TreeStar Inc, Ashland, OR). We used Cytometer Setup & Tracking beads (BD Biosciences) to check for inter-day variability, and Fluorescence Minus One (FMO) controls. We defined MAIT cells as live (DAPI^-^) CD3^+^CD4^-^CD161^hi^Vα7.2^+^ cells, expressed as a percentage of total CD3+ lymphocytes, and used CD38 and CD69 as markers of cell activation.

### Immunohistochemistry

In a separate set of patients, we embedded cryosections from duodenal biopsies in Tissue Tek OCT compound (Sakura USA, Torrance, CA) and used a Leica CM3000 Cryostat (Leica Instruments GmbH, Nussloch, Germany) to cut 5 um sections, picked up on poly-L-lysine coated slides, and air-dried. We stained the sections with primary antibodies against CD3, IL-18Rα, and Vα7.2, followed by corresponding secondary antibodies conjugated to: AF555, AF488, and Cy5. Antibodies were obtained from Dako (Carpinteria, CA), BioLegend, or LifeTechnologies. We counterstained with DAPI to visualize cell nuclei. We used the Nuance Multispectral Imaging system (CRI, Woburn, MA) to visualize and captured images with a digital camera. We analyzed the images with ImageJ software (US National Institutes of Health, Bethesda, MD). We defined MAIT cells as CD3^+^IL-18Rα^+^Vα7.2^+^ cells, as described previously [16].

### Vibriocidal and plasma antibody levels

We performed the vibriocidal assay as previously described [17]. We quantified LPS (prepared from *V. cholerae* O1 as previously described [18])-specific IgA, IgG, and IgM antibody responses in plasma using a kinetic ELISA method [19].

### Markers of intestinal inflammation and permeability

In a subset of patients from whom we had data from both day 2 and day 30, we performed ELISA to determine the concentration of myeloperoxidase (MPO; Alpco, Salem, NH) and alpha anti-trypsin (AAT; ImmuChrom GmBH, NC) from stool samples obtained at the time of admission (day 0), at dilution factors of 1:200 and 1:400, respectively.

### Single Cell TCR sequencing

MAIT cells from PBMCs and LPLs from one donor, at both acute and convalescent stages of infection, were single cell sorted using the Aria II cell sorter (BD Biosciences) directly into One Step RT-PCR reaction mix (NEB) loaded in MicroAmp Optical 96-well reaction plates (Applied Biosystems). MAITs were defined as live (DAPI^-^) CD3^+^CD4^-^CD161^hi^Vα7.2^+^ cells. Following reverse transcription and preamplification reaction, a series of three nested PCR’s were run using primers for TCR sequence and gene expression as described earlier [20]. To separate reads from every well in every plate according to specified barcodes we processed and demultiplexed raw sequencing data using a custom software pipeline described in [20]. The data were analyzed using R package as described [20].

### Statistics

We used the Wilcoxon signed-ranked test for comparisons of frequency and activation of MAIT cells between different study days. We used one-way ANOVA with Tukey’s multiple comparison test for comparison of two or more groups. We log transformed MPO and AAT values and used Spearman’s correlation to determine their association with changes in LPL MAITs. All P values were two-tailed, with a value of <0.05 considered the threshold for statistical significance. We performed analyses using STATA version 13.1 (StataCorp, College Station, TX), and GraphPad Prism version 6.0 (GraphPad Software, Inc., La Jolla, CA).

### Ethics

This study was approved by the Ethical Review and Research Review Committees of the icddr,b, and the Institutional Review Boards of Massachusetts General Hospital and the University of Utah.

## Results

To characterize MAIT cell presence and activity in the duodenum during cholera, we obtained duodenal biopsies from a total of 15 cholera patients, on 10 of whom we performed flow cytometric (FC) analysis and 5 of whom we performed immunohistochemical (IHC) analysis (Supplementary table 1). The median age of the FC group was 31 (range 23 to 36) years, and the median age of the IHC group was 30 (range 26-44) years. Only one female was recruited for each group. All patients mounted at least a 16-fold rise in vibriocidal response by day 7 of illness.

### MAIT cells are found predominantly in the lamina propria of the duodenum, and are more abundant during acute infection than at convalescence

We performed immunohistochemistry from frozen sections of paired duodenal biopsies from five cholera patients. We identified MAIT cells as CD3^+^IL-18Rα^+^Vα7.2^+^ cells (Figure 1A). The majority of MAIT cells were identified in the lamina propria, with no cells identified in the villous and crypt epithelia. By this technique, we found that the frequency of MAIT cells, as a % of total CD3^+^ cells, was significantly higher at day 2 of infection compared to day 30 (\*\**p* = 0.0049, Figure 1B). In contrast, we demonstrated that the occurrence of CD3^+^ cells, as % of total cells in the lamina propria, did not change between acute and convalescent stages of infection (Figure 1C).

**Figure 1.**
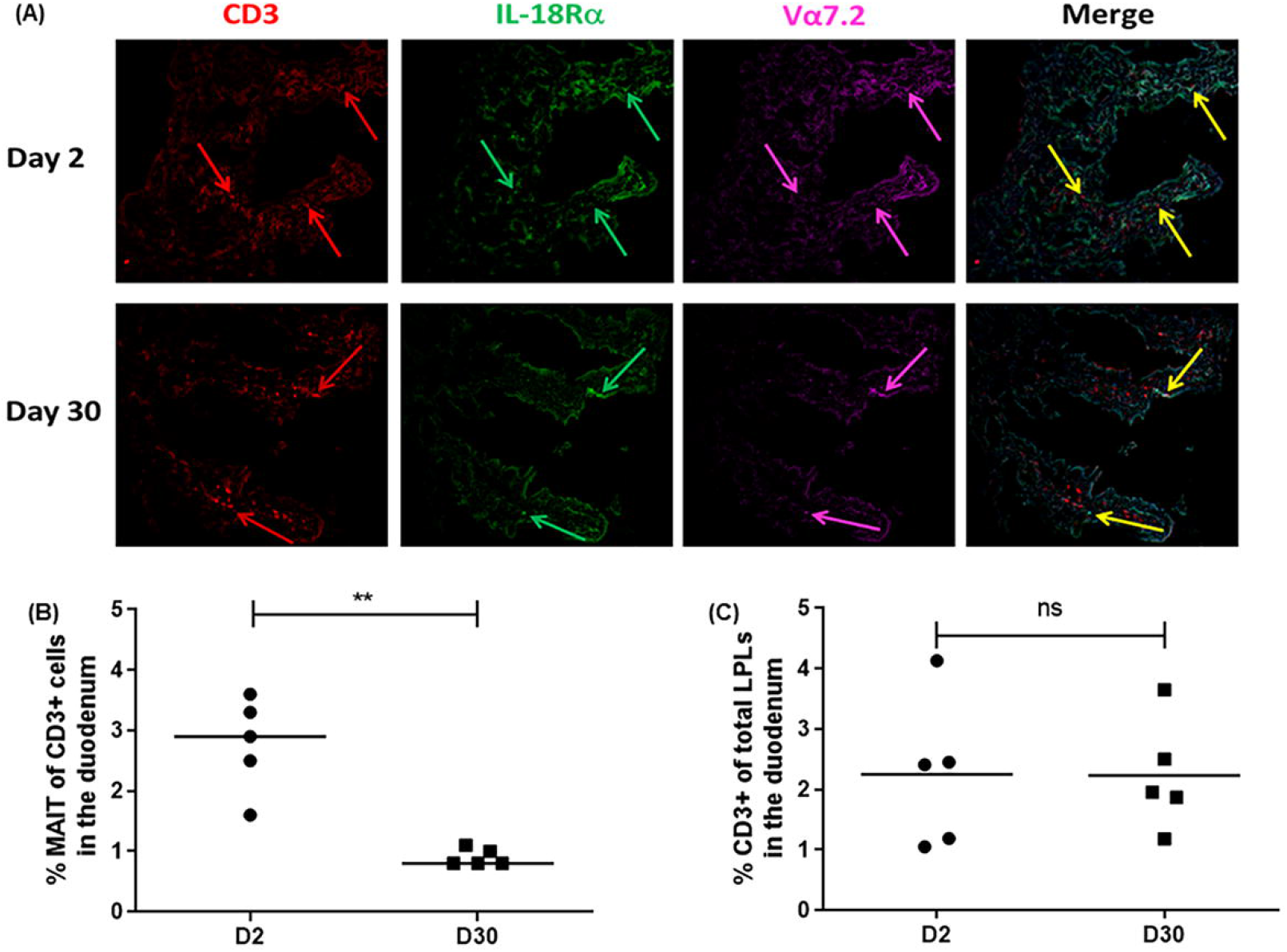
MAIT cell localization and frequency in LP by Immunohistochemistry. Cryosections of duodenal biopsies obtained from five cholera patients were stained with antibodies as mentioned in methods and imaged using Leica CM3000 Cryostat. (A) Representative images of duodenal biopsies at day 2 and day 30, shows CD3^+^ T cells (red arrows), IL-18Rα^+^ T cells (green arrows), Vα 7.2^+^ cells (magenta arrows), and the merged image shows MAIT cells (yellow arrows indicating CD3^+^ IL-18Rα^+^ Vα7.2^+^ cells). (B) Frequency of MAIT cells, as a % of total CD3+ cells in the duodenum at day 2 and day 30 post infection (p.i.), and (C) Frequency of CD3^+^ cells, as % of total cells in the lamina propria at day 2 and day 30 p.i. Statistical significance of the difference between day 2 and day 30 was determined using paired t-test (data passed Shapiro-Wilk normality test). ** denotes *p* ≤ 0.01.

### Compared to peripheral blood MAIT cells, duodenal MAIT cells at both acute and convalescent stages are more activated, but present in similar frequencies

We performed flow cytometric analysis on LPLs isolated from duodenal biopsies in 10 patients, of which only 7 completed 30 days of follow-up. We found that during all phases of cholera, MAIT cells were present in the duodenal lamina propria at frequencies similar to those found in the periphery (Figure 2A). We demonstrated that the occurrence of CD3+ cells, as % of total cells in the lamina propria at the convalescent stage is lower than in the periphery (Figure 2B). Using CD38^+^ as a marker of activation, we found that at both days 2 and 30, duodenal MAIT cells were significantly more activated than peripheral MAIT cells. At day 2, a mean of 60% of all duodenal MAITs were CD38^+^, compared to 15% of all peripheral MAIT cells (95% CI of difference 16.8 – 74.55, P = 0.0005); similarly, at day 30, a mean of 59% of duodenal MAIT cell was activated, compared to 21% of peripheral MAIT cells (95% CI of difference 6 – 70, P < 0.01) (Figure 2C). We also found that at day 30, percentage frequencies of CD69^+^ duodenal MAIT cells were significantly higher than peripheral MAIT cells (P < 0.05) (Figure 2D). We found no significant differences in activation between duodenal MAITs at day 2 and day 30. Figure 2E shows the gating strategy for MAIT cells and representative flow cytometry plots of CD38^+^ and CD69^+^ MAIT cells in the lamina propria and the periphery.

**Figure 2.**
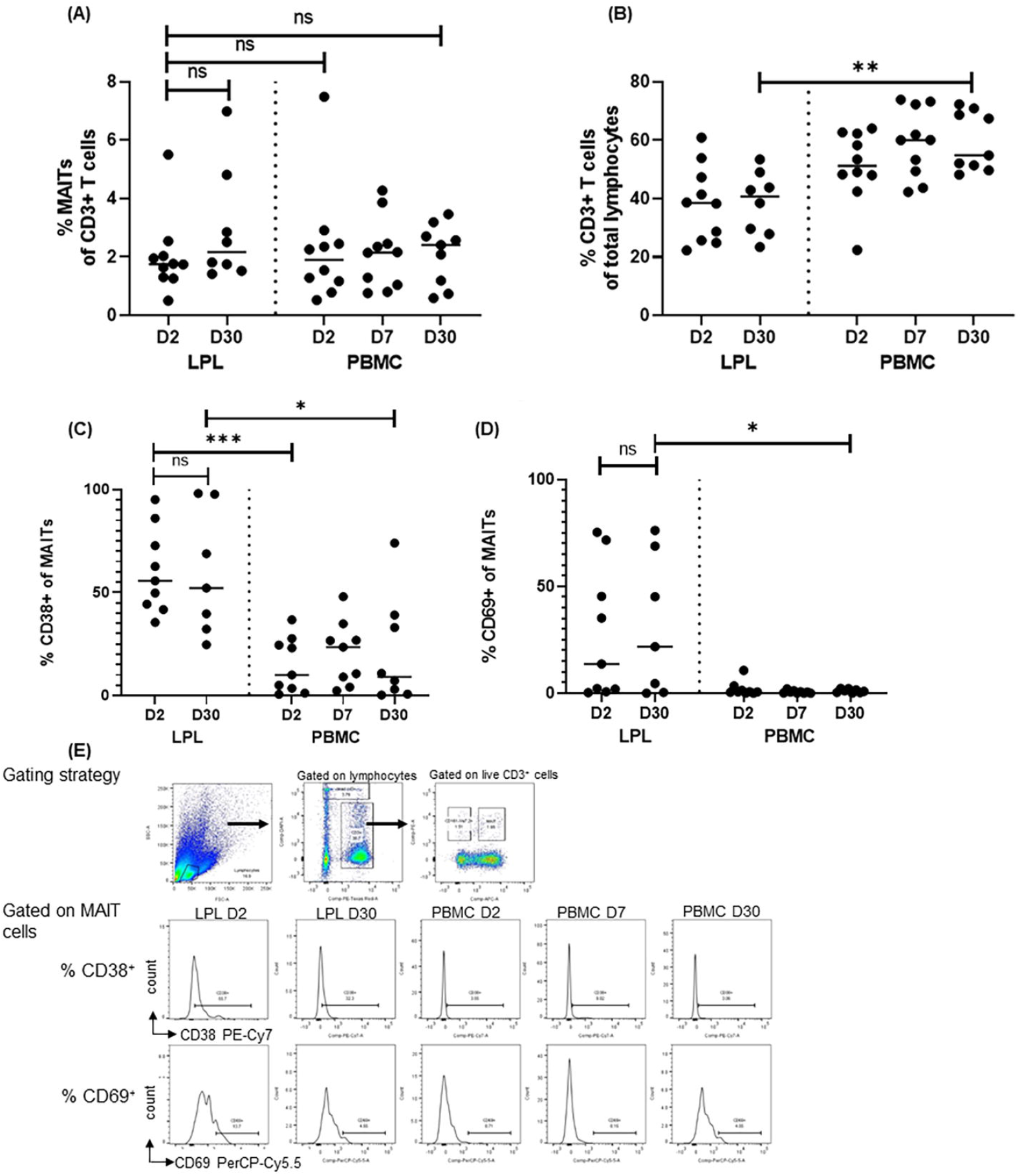
MAIT cell frequency and activation in PBMC and LPL by flow cytometric analysis. Lamina propria lymphocytes (LPLs) were isolated from patients at day 2 and day 30 p.i. and PBMCs were isolated from patients at day 2, day 7, and day 30 p.i. as mentioned in methods. (A) Frequency of MAIT cells (live (DAPI^-^) CD3^+^CD161^hi^Vα7.2^+^ cells), as % of total CD3^+^ lymphocytes in LPL and PBMCs. (B) Frequency of CD3^+^ T cells, as of total lymphocyte population. (C) Frequency of CD38^+^ cells gated on MAIT cells in LPL and PBMCs. (D) Frequency of CD69^+^ cells gated on MAIT cells in LPL and PBMCs. Horizontal bar shows median values in graphs. (E) Gating strategy used to identify MAIT cells and representative histograms showing % CD38+ and % CD69+ cells gated on MAIT cells in LPL and PBMCs. A statistical significantsignificance of difference between groups was determined using one-way ANOVA with Tukey’s post hoc testing. * denotes p ≤ 0.05, ** denotes p ≤ 0.01, and *** denotes p < 0.001.

### Increased intestinal permeability and inflammation are associated with loss of duodenal (but not peripheral) MAIT cells

Given the known cytotoxic capacity of MAIT cells and their activation in inflammatory bowel disease [21, 22], we examined whether baseline intestinal inflammation was associated with changes in MAIT cell frequency observed in IHC. We measured two common fecal markers of intestinal permeability (myeloperoxidase (MPO) and inflammation (alpha-1-1antitrypsin (AAT)) in six of the seven cholera patients from whom we had paired days 2 and 30 MAIT cell data. We found a high level of correlation between the markers and the loss of duodenal MAIT cells from day 2 to 30 (r = 0.90, p = 0.03 for MPO; r = 1.00, P = 0.003 for AAT; Figure 3A and B). We did not find any correlation between these markers and changes in the frequency of peripheral MAIT cells.

**Figure 3.**
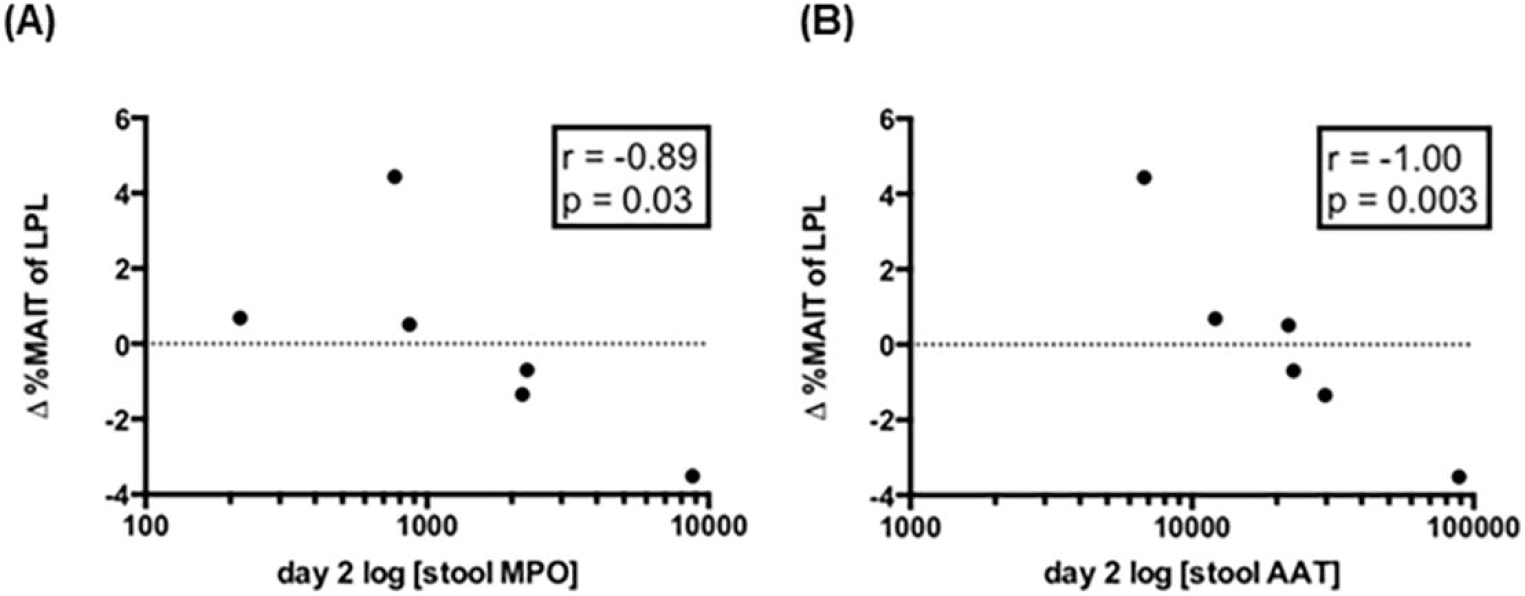
MAIT cell number and correlation with fecal markers of intestinal permeability (myeloperoxidase (MPO)) and inflammation (alpha-1-1antitrypsin (AAT)). Stool markers of intestinal permeability and inflammation were measured using ELISA, values were log transformed and the correlation with LPL MAIT cell number was determined using Spearman’s correlation. (A) shows the correlation of LPL MAITs with MPO and (B) shows the correlation of LPL MAITs with AAT. [δ%MAIT = day30 - day2 %MAIT].

### Circulating MAIT (but not non-MAIT CD3^+^ Vα7.2^-<^) cells from cholera patients express gut-homing markers

To determine the gut-homing potential of circulating MAIT cells, we measured the expression of integrin β7 (which can pair with integrin α4 to form α4β7, a gut-homing marker). We found that at day 7, the majority (mean +/-SD, 65 +/-8.9%) of peripheral MAIT cells in cholera patients expressed β7, compared to non-MAIT cells that expressed β7 in 43% (P < 0.05, Figure 4).

**Figure 4.**
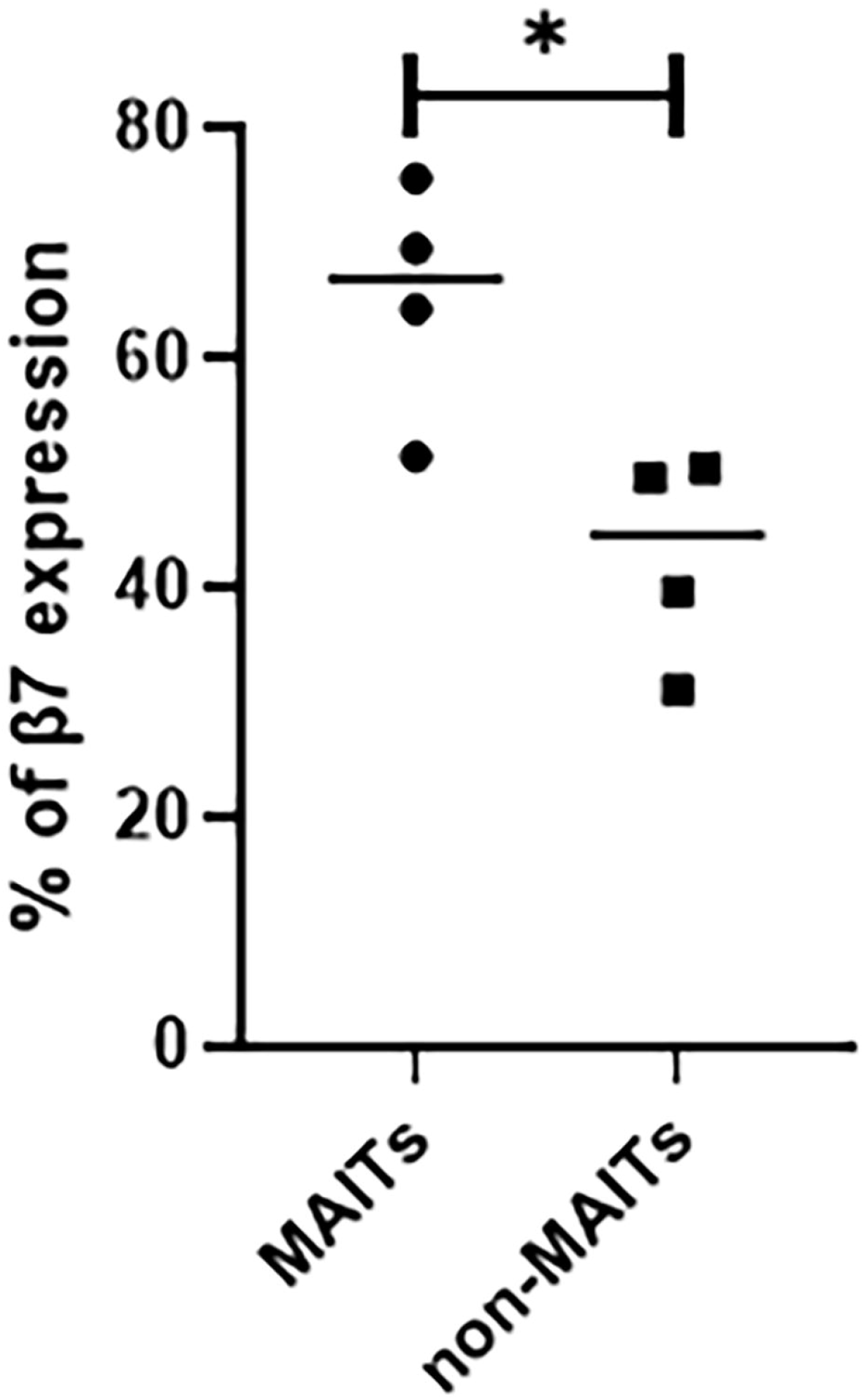
Higher expression of integrin β7 on circulating MAIT cells compared to non-MAIT cells. Expression of β7 was determined on circulating MAIT and non-MAIT (CD3+ Vα7.2-) cells from cholera patients using flow cytometry. Statistical significance of the difference between MAIT and non MAIT cells was determined using unpaired t-test (data passed Shapiro-Wilk normality test). * denotes *p* ≤ 0.05.

### Single-cell TCR analysis of duodenal MAIT cells reveals a different TCR usage than that of MAIT cells in peripheral blood during acute infection

Few studies have reported TCR usage of MAIT cells in tissues [23-25], with minimal data on paired αβ TCR usage. To understand how MAIT TCR repertoire is affected in LPL and PBMCs during cholera, we utilized paired TCR-phenotype single-cell Illumina sequencing as previously described [20], on paired tissue-blood samples from two patients. The paired αβ TCR usage, are shown as heatmaps in Figure 5 (A-D). When we compared TCR usage from PBMC (46 clones sequenced at day 2 and 7) and LPL (46 clones sequenced at day 2) samples that were available during acute infection, we found a dominant MAIT cell clone in the LPL, which was not found in the periphery, expressing TRAV1-2, TRAJ33, TRBV7-2, and TRBJ2-2 (Figure 5A). In addition, we noted that MAIT clones observed in PBMCs obtained at day 7 differed from those observed at day 2 (Figure 5A). When analyzing based on TCR β usage alone, in concurrence with previous studies [23-26], we also found MAIT cells in PBMCs obtained at day 2 and day 7 that expressed TRBV20 and TRBV6 (Figure 5C). In a donor for whom paired PBMC (89 clones sequenced) and LPL (91 clones sequenced) were available at the convalescent stage, we found overlapping MAIT TCR repertoire (Figure 5A), with the majority of PBMC and LPL MAIT cells expressed TRAJ33 (Figure 5B), TRBV7-2, TRBV20-1 or TRBV30 (Figure 5C) and TRBJ 2-2 (Figure 5D).

**Figure 5.**
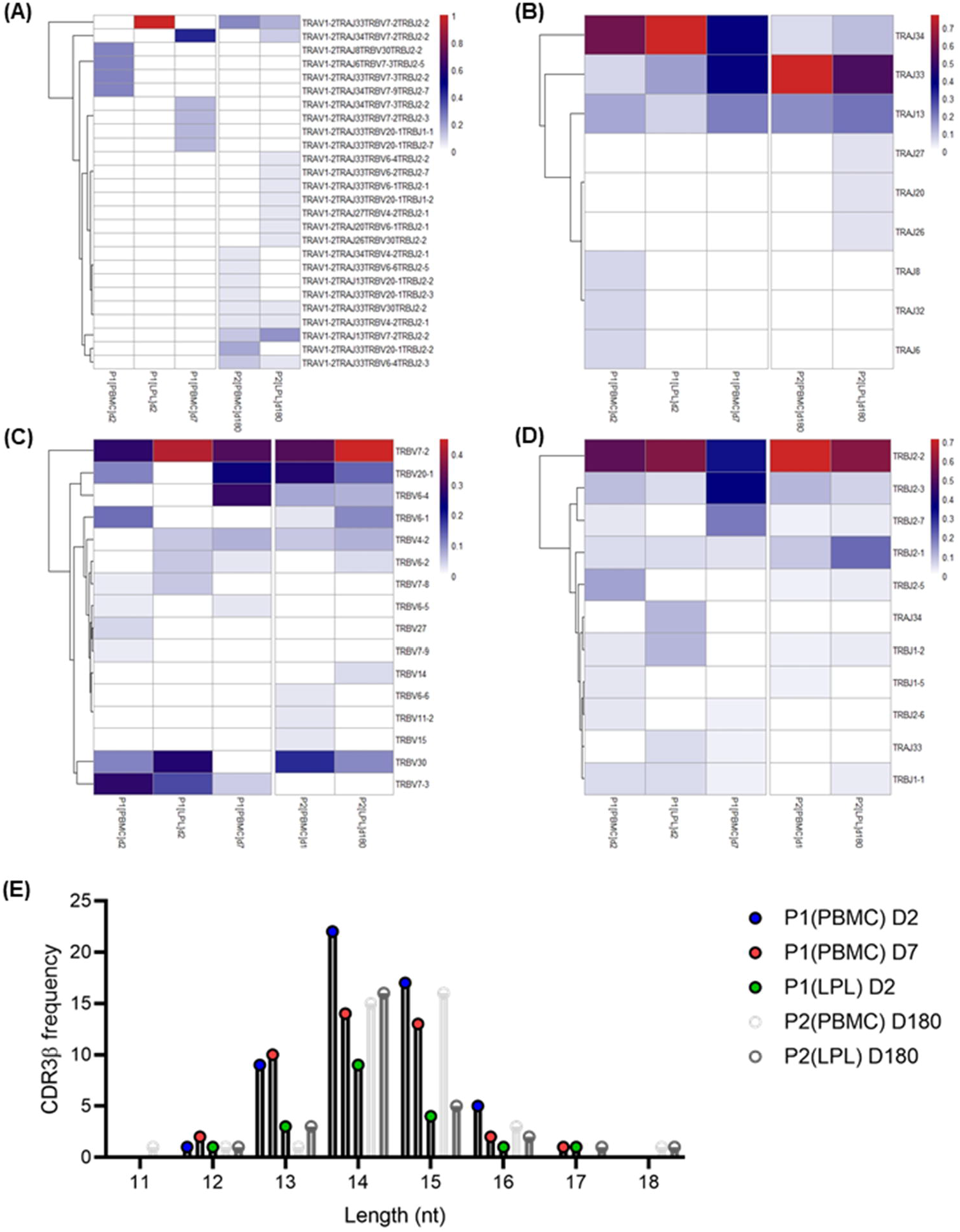
Different distribution of MAIT TCR in LPL compared to PBMCs. MAIT cells were sorted from LPL and PBMCs from two patients at the acute and convalescent stage of illness and TCR usage was analyzed at the single-cell level using Illumina MiSeq sequencing. (A) Paired MAIT TCRαβ usage in each patient is shown as a heat map with hierarchical clustering performed using Euclidean distance. (B, C, and D) TRAJ, TRBV and TRBJ usage in LPL and PBMCs is shown as heat map. (E) The length distribution of MAIT CDR3β sequence in LPL and PBMCs is shown as a bar graph.

MAIT TCR β-chain repertoire diversity resides within the complementarity determining region (CDR) 3β loop [27-29] and to determine if MAIT TCR usage differ at the CDR3 level, we investigated the length distribution of amino acids in the CDR3β region of PBMC and LPL. We observed that the frequency of CDR3β sequences with 13 to 16 nucleotide length was relatively lower in LPL compared to PBMCs at day 2 and comparable at day 180 (Figure 5E). Overall, differential patterns of MAIT TCR usage was observed in the duodenum compared with peripheral blood during acute and convalescent infection.

## Discussion

*V. cholerae* infection is caused by the ingestion of bacteria, followed by colonization of the small intestine where cholera toxin is elaborated, resulting in chloride ion secretion and secretory diarrhea [18]. MAIT cells are innate-like lymphocytes known to provide immediate effector functions in response to infections in human tissues [30, 31]. Although studies have described MAIT cells in the human intestinal mucosa [4, 32-34], there is limited data available on MAIT cells in the intestinal mucosa during an acute intestinal infection. In this study, we performed endoscopy and obtained duodenal biopsies in a cohort of patients with culture-confirmed severe dehydrating *V. cholerae* O1 infection. We showed that during acute human cholera, MAIT cells in the duodenal mucosa are present at frequencies similar to that seen in the peripheral circulation. Using immunohistochemistry and multispectral imaging, we demonstrated that the vast majority of MAIT cells are in the lamina propria, predominantly in the crypts, and rarely in the epithelia. This is consistent with a previous report using MR1 tetramer staining of healthy human jejunal tissue, showing that MAIT cells reside predominantly near the base of the villi [35].

We have previously shown, in endoscopically-obtained biopsies from cholera patients, that during acute disease, there is an upregulation of innate responses, including infiltration of neutrophils, degranulation of mast cells, and expression of pro-inflammatory cytokines [36, 37]. Using immunohistochemistry here, we showed that MAIT cell frequencies are significantly increased during acute infection compared to convalescence, although we were unable to replicate this finding using flow cytometric analysis of LPLs, which used different markers to identify MAIT cells. We have previously shown that in adults with cholera, peripheral MAIT cells are highly activated at day 7 following infection, and that MAIT frequencies remain stable for up to 90 days following infection [14]. Reports of MAIT cell kinetics in the intestinal mucosa are lacking, although studies have shown that MAIT cells are present and active in the gastric mucosa during *H. pylori* infection [38, 39], at decreased frequencies in duodenal lamina propria in celiac disease patients compared to healthy controls [32], and decreased in the colon in chronic HIV infection [40, 41]. It was recently reported that MAIT cells are decreased in the circulation and accumulate in the inflamed mucosa of patients with inflammatory bowel diseases (IBD), where they display increased cytokine secretion capacities [16, 34, 42, 43]. In this study, we showed that during cholera, an acute bacterial enteric infection, MAIT cells in the duodenum are activated at levels significantly higher than that in the peripheral blood. Taken together, MAIT cells are highly activated and present in the lamina propria of the duodenum during cholera. We hypothesize that they play an important role in the innate immune response to cholera.

Studies in humans with celiac disease, characterized by increased small bowel permeability, have shown an association between intestinal pathology and loss of intestinal MAIT cells [32]. Similarly, studies of ileal biopsies from patients with inflammatory bowel disease showed an accumulation of MAIT cells in inflamed compared to healthy tissue [16]. We hypothesized that compromised gut barrier function would increase MAIT cell exposure to microbes, resulting in MAIT cell activation and cell death. Thus, we examined two common fecal markers of intestinal inflammation (MPO) and permeability (AAT) [44], and found a high heterogeneity among cholera patients. Notably, we showed that levels of these markers were highly correlated with the loss of duodenal, but not peripheral, MAIT cells. While these findings suggest that MAIT cell loss is associated with intestinal inflammation and permeability, we cannot make conclusions regarding causality or pathogenesis.

Recent studies suggest that variability in MAIT TCR affects microbial ligand discrimination, activation, and phenotype [45-47]. In our TCR analysis of two donors, we observed that during acute infection (P1) there was a different distribution of MAIT clones in LPL compared to PBMCs. However, in the convalescent stage (P2), there was overlapping utilization of TCR usage. We hypothesize from these observations that during acute infection, the intestinal compartment may have more layers of functional and phenotypic heterogeneity of MAIT cells than seen in the peripheral blood. The low sample size and lack of paired blood and LPL samples significantly limit our TCR analyses, and thus further studies would be needed to confirm our observations regarding the TCR usage between LPL and blood MAITs. One of the limitations of our study is that we were not able to assess cytokine secretion of MAIT cells during acute and convalescent stages of infection and further investigation of MAIT cell functionality in cholera is warranted.

In conclusion, we have shown that MAIT cells are present in the lamina propria of the duodenum and are highly activated compared to peripheral blood during human cholera infection. We showed lower frequencies of activated MAITs and higher β7 expression in peripheral blood suggesting their gut-homing potential. Additional studies exploring the functional cytotoxic abilities of duodenal MAIT cells to inactivate *V. cholerae* or their roles in innate and adaptive immune responses are needed.

## Data Availability

All relevant data are within the manuscript and its Supporting Information files.

## Acknowledgments

We thank the patients for participating in this study and the field workers and research staff at the icddr,b, Bangladesh, for their support and effort in sample collection and processing.

This work was supported in part by core grants to the icddr,b and by the Government of the People’s Republic of Bangladesh, Global Affairs Canada (GAC), Swedish International Development Cooperation Agency (SIDA), and the Department of International Development (UKAid). This work was also supported by grants from the National Institutes of Health (U01AI058935 to E.T.R.; R01AI135115 to D.T.L. and F.Q.; R01AI130378 to D.T.L. and T.R.B.), the Fogarty International Center (FIC) and NIAID training grant in vaccine development and public health (TW005572 to M.H. and T.R.B.), and an FIC Global Emerging Leader Award (K43TW010362 to T.R.B.), and a Thrasher Research Fund Early Career Award [D.T.L.]. The funders had no role in study design, data collection and interpretation, or the decision to submit the work for publication.

## References

1. Le Bourhis L, Guerri L, Dusseaux M, Martin E, Soudais C, Lantz O. Mucosal-associated invariant T cells: unconventional development and function. Trends in immunology. 2011;32(5):212–8. doi: 10.1016/j.it.2011.02.005. PubMed PMID: 21459674.

2. Ussher JE, Klenerman P, Willberg CB. Mucosal-associated invariant T-cells: new players in anti-bacterial immunity. Frontiers in immunology. 2014;5:450. doi: 10.3389/fimmu.2014.00450. PubMed PMID: 25339949; PubMed Central PMCID: PMC4189401.

3. Dusseaux M, Martin E, Serriari N, Peguillet I, Premel V, Louis D, et al. Human MAIT cells are xenobiotic-resistant, tissue-targeted, CD161hi IL-17-secreting T cells. Blood. 2011;117(4):1250–9. Epub 2010/11/19. doi: 10.1182/blood-2010-08-303339. PubMed PMID: 21084709.

4. Reantragoon R, Corbett AJ, Sakala IG, Gherardin NA, Furness JB, Chen Z, et al. Antigen-loaded MR1 tetramers define T cell receptor heterogeneity in mucosal-associated invariant T cells. J Exp Med. 2013;210(11):2305–20. Epub 2013/10/09. doi: 10.1084/jem.20130958. PubMed PMID: 24101382; PubMed Central PMCID: PMCPMC3804952.

5. Corbett AJ, Eckle SB, Birkinshaw RW, Liu L, Patel O, Mahony J, et al. T-cell activation by transitory neo-antigens derived from distinct microbial pathways. Nature. 2014;509(7500):361–5. doi: 10.1038/nature13160. PubMed PMID: 24695216.

6. McWilliam HE, Birkinshaw RW, Villadangos JA, McCluskey J, Rossjohn J. MR1 presentation of vitamin B-based metabolite ligands. Current opinion in immunology. 2015;34:28–34. doi: 10.1016/j.coi.2014.12.004. PubMed PMID: 25603223.

7. Le Bourhis L, Dusseaux M, Bohineust A, Bessoles S, Martin E, Premel V, et al. MAIT Cells Detect and Efficiently Lyse Bacterially-Infected Epithelial Cells. PLoS pathogens. 2013;9(10):e1003681. doi: 10.1371/journal.ppat.1003681. PubMed PMID: 24130485; PubMed Central PMCID: PMC3795036.

8. Kurioka A, Ussher JE, Cosgrove C, Clough C, Fergusson JR, Smith K, et al. MAIT cells are licensed through granzyme exchange to kill bacterially sensitized targets. Mucosal Immunol. 2015;8(2):429–40. Epub 2014/10/02. doi: 10.1038/mi.2014.81. PubMed PMID: 25269706; PubMed Central PMCID: PMCPMC4288950.

9. Ali M, Nelson AR, Lopez AL, Sack DA. Updated global burden of cholera in endemic countries. PLoS neglected tropical diseases. 2015;9(6):e0003832. doi: 10.1371/journal.pntd.0003832. PubMed PMID: 26043000; PubMed Central PMCID: PMC4455997.

10. Bhuiyan TR, Lundin SB, Khan AI, Lundgren A, Harris JB, Calderwood SB, et al. Cholera caused by Vibrio cholerae O1 induces T-cell responses in the circulation. Infection and immunity. 2009;77(5):1888–93. Epub 2009/02/23. doi: 10.1128/IAI.01101-08. PubMed PMID: 19237532.

11. Harris AM, Bhuiyan MS, Chowdhury F, Khan AI, Hossain A, Kendall EA, et al. Antigen-Specific Memory B-Cell Responses to <em>Vibrio cholerae</em> O1 Infection in Bangladesh. Infection and Immunity. 2009;77(9):3850–6. doi: 10.1128/iai.00369-09.

12. Qadri F, Raqib R, Ahmed F, Rahman T, Wenneras C, Das SK, et al. Increased levels of inflammatory mediators in children and adults infected with Vibrio cholerae O1 and O139. Clinical and diagnostic laboratory immunology. 2002;9(2):221–9. PubMed PMID: 11874856; PubMed Central PMCID: PMC119937.

13. Qadri F, Bhuiyan TR, Dutta KK, Raqib R, Alam MS, Alam NH, et al. Acute dehydrating disease caused by Vibrio cholerae serogroups O1 and O139 induce increases in innate cells and inflammatory mediators at the mucosal surface of the gut. Gut. 2004;53(1):62–9. PubMed PMID: 14684578; PubMed Central PMCID: PMC1773936.

14. Leung DT, Bhuiyan TR, Nishat NS, Hoq MR, Aktar A, Rahman MA, et al. Circulating mucosal associated invariant T cells are activated in Vibrio cholerae O1 infection and associated with lipopolysaccharide antibody responses. PLoS Negl Trop Dis. 2014;8(8):e3076. Epub 2014/08/22. doi: 10.1371/journal.pntd.0003076. PubMed PMID: 25144724; PubMed Central PMCID: PMCPMC4140671.

15. Uddin T, Harris JB, Bhuiyan TR, Shirin T, Uddin MI, Khan AI, et al. Mucosal immunologic responses in cholera patients in Bangladesh. Clinical and vaccine immunology : CVI. 2011;18(3):506–12. doi: 10.1128/CVI.00481-10. PubMed PMID: 21248157; PubMed Central PMCID: PMC3067383.

16. Serriari NE, Eoche M, Lamotte L, Lion J, Fumery M, Marcelo P, et al. Innate mucosal-associated invariant T (MAIT) cells are activated in inflammatory bowel diseases. Clin Exp Immunol. 2014;176(2):266–74. Epub 2014/01/24. doi: 10.1111/cei.12277. PubMed PMID: 24450998; PubMed Central PMCID: PMCPMC3992039.

17. Johnson RA, Uddin T, Aktar A, Mohasin M, Alam MM, Chowdhury F, et al. Comparison of immune responses to the O-specific polysaccharide and lipopolysaccharide of Vibrio cholerae O1 in Bangladeshi adult patients with cholera. Clinical and vaccine immunology : CVI. 2012;19(11):1712–21. Epub 2012/09/19. doi: 10.1128/CVI.00321-12. PubMed PMID: 22993410.

18. Johnson RA, Uddin T, Aktar A, Mohasin M, Alam MM, Chowdhury F, et al. Comparison of immune responses to the O-specific polysaccharide and lipopolysaccharide of Vibrio cholerae O1 in Bangladeshi adult patients with cholera. Clin Vaccine Immunol. 2012;19(11):1712–21. Epub 2012/09/21. doi: 10.1128/CVI.00321-12. PubMed PMID: 22993410; PubMed Central PMCID: PMCPMC3491541.

19. Qadri F, Ahmed F, Karim MM, Wenneras C, Begum YA, Abdus Salam M, et al. Lipopolysaccharide-and cholera toxin-specific subclass distribution of B-cell responses in cholera. Clinical and diagnostic laboratory immunology. 1999;6(6):812–8. PubMed PMID: 10548569; PubMed Central PMCID: PMC95781.

20. Han A, Glanville J, Hansmann L, Davis MM. Linking T-cell receptor sequence to functional phenotype at the single-cell level. Nature Biotechnology. 2014;32(7):684–92. doi: 10.1038/nbt.2938.

21. Ju JK, Cho YN, Park KJ, Kwak HD, Jin HM, Park SY, et al. Activation, Deficiency, and Reduced IFN-γ Production of Mucosal-Associated Invariant T Cells in Patients with Inflammatory Bowel Disease. Journal of Innate Immunity. 2020;12(5):422–34. doi: 10.1159/000507931.

22. Serriari N-E, Eoche M, Lamotte L, Lion J, Fumery M, Marcelo P, et al. Innate mucosal-associated invariant T (MAIT) cells are activated in inflammatory bowel diseases. Clinical & Experimental Immunology. 2014;176(2):266-74. doi: https://doi.org/10.1111/cei.12277.

23. Loh L, Gherardin NA, Sant S, Grzelak L, Crawford JC, Bird NL, et al. Human Mucosal-Associated Invariant T Cells in Older Individuals Display Expanded TCRαβ Clonotypes with Potent Antimicrobial Responses. The Journal of Immunology. 2020;204(5):1119–33. doi: 10.4049/jimmunol.1900774.

24. Voillet V, Buggert M, Slichter CK, Berkson JD, Mair F, Addison MM, et al. Human MAIT cells exit peripheral tissues and recirculate via lymph in steady state conditions. JCI Insight. 2018;3(7):e98487. doi: 10.1172/jci.insight.98487. PubMed PMID: 29618662.

25. Wong EB, Gold MC, Meermeier EW, Xulu BZ, Khuzwayo S, Sullivan ZA, et al. TRAV1-2+ CD8+ T-cells including oligoconal expansions of MAIT cells are enriched in the airways in human tuberculosis. Communications Biology. 2019;2(1):203. doi: 10.1038/s42003-019-0442-2.

26. Lepore M, Kalinichenko A, Colone A, Paleja B, Singhal A, Tschumi A, et al. Parallel T-cell cloning and deep sequencing of human MAIT cells reveal stable oligoclonal TCRβ repertoire. Nat Commun. 2014;5:3866. Epub 2014/05/17. doi: 10.1038/ncomms4866. PubMed PMID: 24832684.

27. Gherardin Nicholas A, Keller Andrew N, Woolley Rachel E, Le Nours J, Ritchie David S, Neeson Paul J, et al. Diversity of T Cells Restricted by the MHC Class I-Related Molecule MR1 Facilitates Differential Antigen Recognition. Immunity. 2016;44(1):32-45. doi: https://doi.org/10.1016/j.immuni.2015.12.005.

28. Gold MC, McLaren JE, Reistetter JA, Smyk-Pearson S, Ladell K, Swarbrick GM, et al. MR1-restricted MAIT cells display ligand discrimination and pathogen selectivity through distinct T cell receptor usage. The Journal of Experimental Medicine. 2014;211(8):1601–10. doi: 10.1084/jem.20140507.

29. Howson LJ, Napolitani G, Shepherd D, Ghadbane H, Kurupati P, Preciado-Llanes L, et al. MAIT cell clonal expansion and TCR repertoire shaping in human volunteers challenged with Salmonella Paratyphi A. Nature Communications. 2018;9(1). doi: 10.1038/s41467-017-02540-x.

30. Amini A, Pang D, Hackstein C-P, Klenerman P. MAIT Cells in Barrier Tissues: Lessons from Immediate Neighbors. Frontiers in immunology. 2020;11(3040). doi: 10.3389/fimmu.2020.584521.

31. Kurioka A, Ussher JE, Cosgrove C, Clough C, Fergusson JR, Smith K, et al. MAIT cells are licensed through granzyme exchange to kill bacterially sensitized targets. Mucosal immunology. 2015;8(2):429–40. doi: 10.1038/mi.2014.81.

32. Dunne MR, Elliott L, Hussey S, Mahmud N, Kelly J, Doherty DG, et al. Persistent changes in circulating and intestinal gammadelta T cell subsets, invariant natural killer T cells and mucosal-associated invariant T cells in children and adults with coeliac disease. PloS one. 2013;8(10):e76008. doi: 10.1371/journal.pone.0076008. PubMed PMID: 24124528; PubMed Central PMCID: PMC3790827.

33. Dunne MR, Elliott L, Hussey S, Mahmud N, Kelly J, Doherty DG, et al. Persistent Changes in Circulating and Intestinal γδ T Cell Subsets, Invariant Natural Killer T Cells and Mucosal-Associated Invariant T Cells in Children and Adults with Coeliac Disease. PloS one. 2013;8(10):e76008. doi: 10.1371/journal.pone.0076008.

34. Haga K, Chiba A, Shibuya T, Osada T, Ishikawa D, Kodani T, et al. MAIT cells are activated and accumulated in the inflamed mucosa of ulcerative colitis. J Gastroenterol Hepatol. 2016;31(5):965–72. Epub 2015/11/22. doi: 10.1111/jgh.13242. PubMed PMID: 26590105.

35. Patel O, Kjer-Nielsen L, Le Nours J, Eckle SB, Birkinshaw R, Beddoe T, et al. Recognition of vitamin B metabolites by mucosal-associated invariant T cells. Nat Commun. 2013;4:2142. Epub 2013/07/13. doi: 10.1038/ncomms3142. PubMed PMID: 23846752.

36. Kuchta A, Rahman T, Sennott EL, Bhuyian TR, Uddin T, Rashu R, et al. <span class=“named-content genus-species” id=“named-content-1”>Vibrio cholerae</span> O1 Infection Induces Proinflammatory CD4<sup>+</sup> T-Cell Responses in Blood and Intestinal Mucosa of Infected Humans. Clinical and Vaccine Immunology. 2011;18(8):1371–7. doi: 10.1128/cvi.05088-11.

37. Qadri F, Bhuiyan TR, Dutta KK, Raqib R, Alam MS, Alam NH, et al. Acute dehydrating disease caused by <em>Vibrio cholerae</em> serogroups O1 and O139 induce increases in innate cells and inflammatory mediators at the mucosal surface of the gut. Gut. 2004;53(1):62–9. doi: 10.1136/gut.53.1.62.

38. Booth JS, Salerno-Goncalves R, Blanchard TG, Patil SA, Kader HA, Safta AM, et al. Mucosal-Associated Invariant T Cells in the Human Gastric Mucosa and Blood: Role in Helicobacter pylori Infection. Frontiers in immunology. 2015;6:466. doi: 10.3389/fimmu.2015.00466. PubMed PMID: 26441971; PubMed Central PMCID: PMC4585133.

39. Booth JS, Salerno-Goncalves R, Blanchard TG, Patil SA, Kader HA, Safta AM, et al. Mucosal-Associated Invariant T Cells in the Human Gastric Mucosa and Blood: Role in Helicobacter pylori Infection. Frontiers in immunology. 2015;6(466). doi: 10.3389/fimmu.2015.00466.

40. Cosgrove C, Ussher JE, Rauch A, Gärtner K, Kurioka A, Hühn MH, et al. Early and nonreversible decrease of CD161++/MAIT cells in HIV infection. Blood. 2013;121(6):951–61. doi: 10.1182/blood-2012-06-436436.

41. Leeansyah E, Ganesh A, Quigley MF, Sonnerborg A, Andersson J, Hunt PW, et al. Activation, exhaustion, and persistent decline of the antimicrobial MR1-restricted MAIT-cell population in chronic HIV-1 infection. Blood. 2013;121(7):1124–35. doi: 10.1182/blood-2012-07-445429.

42. Hiejima E, Kawai T, Nakase H, Tsuruyama T, Morimoto T, Yasumi T, et al. Reduced Numbers and Proapoptotic Features of Mucosal-associated Invariant T Cells as a Characteristic Finding in Patients with Inflammatory Bowel Disease. Inflamm Bowel Dis. 2015;21(7):1529–40. Epub 2015/05/07. doi: 10.1097/mib.0000000000000397. PubMed PMID: 25946569.

43. Tominaga K, Yamagiwa S, Setsu T, Kimura N, Honda H, Kamimura H, et al. Possible involvement of mucosal-associated invariant T cells in the progression of inflammatory bowel diseases. Biomed Res. 2017;38(2):111–21. Epub 2017/04/27. doi: 10.2220/biomedres.38.111. PubMed PMID: 28442662.

44. Keusch GT, Denno DM, Black RE, Duggan C, Guerrant RL, Lavery JV, et al. Environmental enteric dysfunction: pathogenesis, diagnosis, and clinical consequences. Clinical infectious diseases : an official publication of the Infectious Diseases Society of America. 2014;59 Suppl 4:S207–12. doi: 10.1093/cid/ciu485. PubMed PMID: 25305288; PubMed Central PMCID: PMC4481570.

45. Dias J, Leeansyah E, Sandberg JK. Multiple layers of heterogeneity and subset diversity in human MAIT cell responses to distinct microorganisms and to innate cytokines. Proceedings of the National Academy of Sciences. 2017;114(27):E5434–E43. doi: 10.1073/pnas.1705759114.

46. Howson LJ, Napolitani G, Shepherd D, Ghadbane H, Kurupati P, Preciado-Llanes L, et al. MAIT cell clonal expansion and TCR repertoire shaping in human volunteers challenged with Salmonella Paratyphi A. Nature Communications. 2018;9(1):253. doi: 10.1038/s41467-017-02540-x.

47. Narayanan GA, McLaren JE, Meermeier EW, Ladell K, Swarbrick GM, Price DA, et al. The MAIT TCRβ chain contributes to discrimination of microbial ligand. Immunology & Cell Biology. 2020;98(9):770-81. doi: https://doi.org/10.1111/imcb.12370.

