## Supplemental Table 1 for "Mucosal-Associated Invariant T (MAIT) Cells are Highly Activated in Duodenal Tissue of Humans with *Vibrio cholerae* O1 Infection"

**Supplementary table 1** Demographics and vibriocidal antibody responses of study subjects.

| | | | | Vibriocidal titer | | | | | |
|---|---|---|---|---|---|---|---|---|---|
| | | | | Ogawa | | | Inaba | | |
| **Subject ID** | **Gender** | **Age range** | **Blood group** | **D2** | **D7** | **D30** | **D2** | **D7** | **D30** |
| SEGD 17 | M | 18-30 | O+VE | 160 | 2560 | 320 | 40 | 1280 | 80 |
| SEGD 18 | M | 18-30 | O+VE | 80 | 1280 | 320 | 10 | 80 | 40 |
| SEGD 19 | M | 31-40 | B+VE | 160 | 5120 | 1280 | 5 | 40 | 20 |
| SEGD 20 | M | 31-40 | O+VE | 5 | 640 | 80 | 5 | 40 | 10 |
| SEGD 21 | M | 31-40 | A+VE | 10 | 1280 | 640 | 5 | 40 | 20 |
| SEGD 22 | M | 18-30 | A+VE | 5 | 640 | 320 | 5 | 320 | 320 |
| SEGD 23 | M | 31-40 | O+VE | 20 | 1280 | 640 | 80 | 640 | 640 |
| SEGD 24 | M | 18-30 | A+VE | 40 | 2560 | 1280 | 10 | 1280 | 1280 |
| SEGD 25 | M | 31-40 | O+VE | 10 | 2560 | Drop out | 40 | 5120 | Drop out |
| SEGD 26 | F | 31-40 | O+VE | 5 | 5120 | 640 | 5 | 2560 | 160 |
| PIC45 | F | 41-50 | B+VE | 5 | 5120 | 1280 | 5 | 5120 | 2560 |
| PIC51 | M | 31-40 | B+VE | 320 | 5120 | 2560 | 320 | 2560 | 1280 |
| PIC53 | M | 18-30 | A+VE | 1280 | 10240 | 1280 | 2560 | 5120 | 2560 |
| PIC94 | M | 18-30 | A+VE | 5 | 5120 | 2560 | 80 | 80 | 80 |
| PIC99 | M | 18-30 | O+VE | 320 | 10240 | 2560 | 640 | 5120 | 2560 |
